# Unsupervised feature extraction using deep learning empowers discovery of genetic determinants of the electrocardiogram

**DOI:** 10.1101/2024.10.07.24314993

**Authors:** Ewa Sieliwonczyk, Arunashis Sau, Konstantinos Patlatzoglou, Kathryn A. McGurk, Libor Pastika, Prisca K Thami, Massimo Mangino, Sean L Zheng, George Powell, Lara Curran, Rachel J Buchan, Pantazis Theotokis, Nicholas S Peters, Bart Loeys, Daniel B. Kramer, Jonathan W Waks, Fu Siong Ng, James S Ware

## Abstract

Advanced data-driven methods can outperform conventional features in electrocardiogram (ECG) analysis, but often lack interpretability. The variational autoencoder (VAE), a form of unsupervised machine learning, can address this shortcoming by extracting comprehensive and interpretable new ECG features. Our novel VAE model, trained on a dataset comprising over one million secondary care median beat ECGs, and validated using the UK Biobank, reveals 20 independent features that capture ECG information content with high reconstruction accuracy. Through phenome- and genome-wide association studies, we illustrate the increased power of the VAE approach for gene discovery, compared with conventional ECG traits, and identify previously unrecognised common and rare variant determinants of ECG morphology. Additionally, to highlight the interpretability of the model, we provide detailed visualisation of the associated ECG alterations. Our study shows that the VAE provides a valuable tool for advancing our understanding of cardiac function and its genetic underpinnings.

## Introduction

The application of machine learning techniques to understand cardiac electrophysiology has sharply risen in recent years, particularly in their application to the electrocardiogram (ECG)^1,2^. Many of these applications rely on supervised models, which are designed to identify specific pre-defined diagnoses or predict outcomes based on the raw ECG signal as input. Apart from features directly related to cardiac function, machine learning-based models have been remarkably successful at deducing more general characteristics from the ECG, such as age and BMI^3,4,5^.

Despite their impressive performance, the inner workings of these models are obscure, contributing to their reputation as a “black box”. This lack of transparency and interpretability limits the potential to generate novel biological insights. More interpretable unsupervised models, such as the variational autoencoder (VAE), have been applied to the ECG^6^ as an alternative approach.

The VAE combines principles derived from machine learning and Bayesian inference to deconstruct a signal into a limited number of highly informative features, also called latent factors (LF). These features are optimized to capture the extent of inter-sample variability and can be used for signal generation^7^, denoising^8^ or predictive and diagnostic models^6^. The features extracted by the VAE are interpretable and lend themselves easily to visualization, enhancing the model’s transparency and facilitating a deeper exploration of the underlying data dynamics.

Most conventional human-defined ECG features have emerged as patterns that are easy for human observers to recognise and label, and then correlate with clinical significance. By contrast, VAE latent factors are optimised to maximise the capture of data content in the ECG, without being constrained by human optical recognition or by prior knowledge and biases. We hypothesised that analysis of the genetic and phenotypic associations of this optimised feature set would empower discovery when compared with analysis of conventional ECG features, and therefore yield novel insights into cardiac electrophysiology and related diseases.

## Results

### Accurate median ECG reconstructions with 20 novel independent ECG features

The VAE was trained on 1,048,778 median ECG beats derived from resting 12-lead ECGs of patients examined at the Beth Israel Deaconess Medical Center (BIDMC), a secondary care centre in Boston, USA. Eight leads were analysed - the first two limb leads and six chest leads. Lead III and the augmented limb leads were not used as they are linear combinations of leads I and II and therefore provide no additional information. Hyperparameter tuning was performed using the validation set (n = 58,265 ECGs) and performance was evaluated in the test set (n = 58,268 ECGs). External validation was performed on digital ECG recordings from the UK Biobank (UKB) (n = 42,248). Median beats were extracted using the Beth Israel Analysis of Vectors of the Heart (BRAVEHEART) software as previously described.^9^ Further details on these datasets are available in the Supplementary Materials.

When comparing the reconstructions to the original medians, we observed an excellent mean Pearson correlation coefficient of 0.94 (±0.11 SD) in the test set and 0.95 (±0.12 SD) in the UKB. As demonstrated in Fig. 1A, the reconstructions accurately captured the typical ECG morphology.

**Figure 1:**
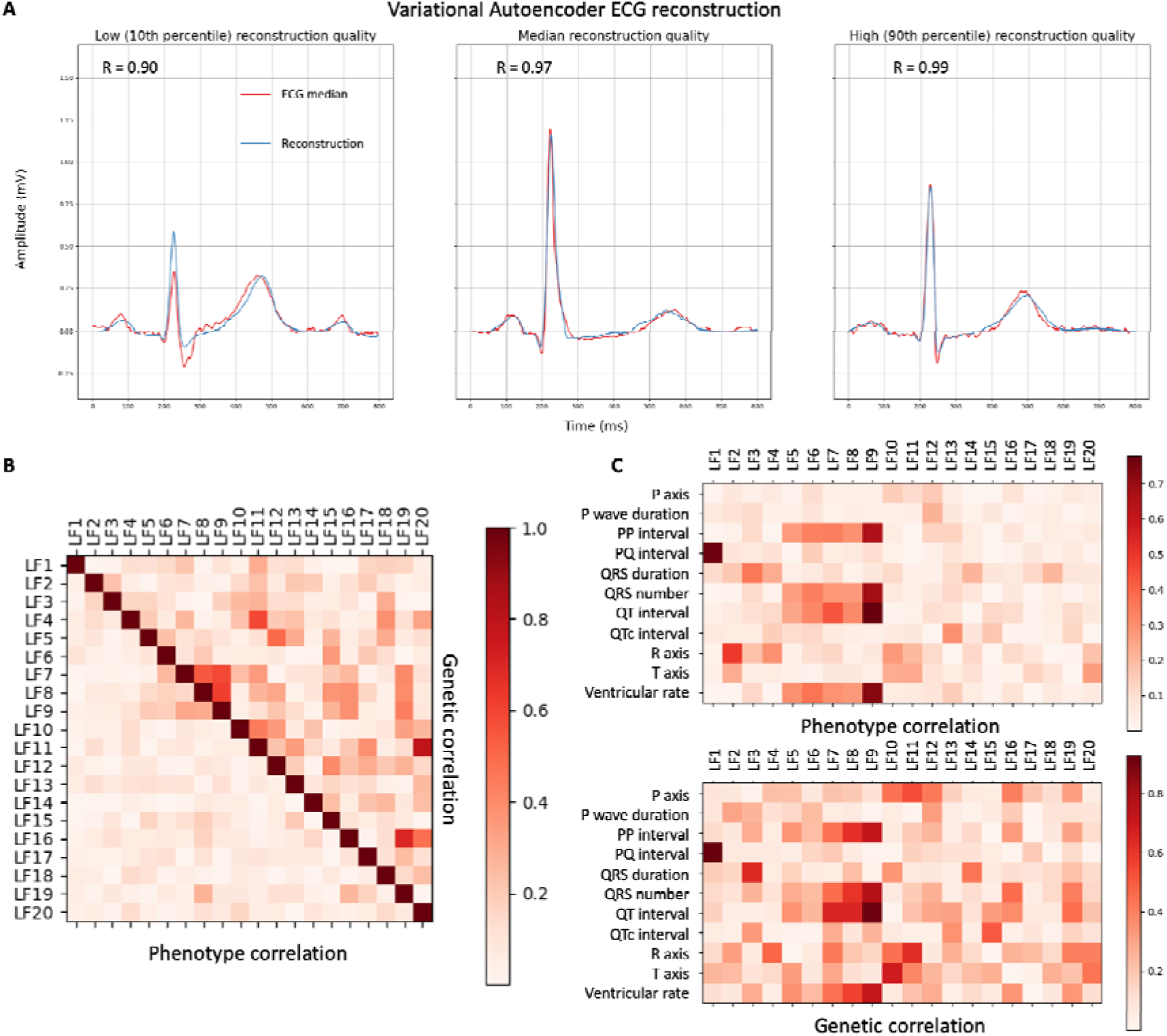
Variational Autoencoder Accuracy and Latent Factor Correlations. 1A: The Variational Autoencoder produces precise median beat reconstructions (blue) that closely match the original ECG medians (red). Example reconstructions spanning a range of accuracies are shown, by sampling reconstructions at the 10th percentile, median, and 90th percentile of the correlation distribution. 1B: A heatmap illustrates the absolute phenotypical correlations (Pearson’s R, bottom left) and absolute pairwise genetic correlations (rg, SNP-based heritability, top right) between individual latent factors, indicating phenotypically uncorrelated novel ECG features with a shared genetic architecture. 1C: The heatmaps depict the absolute phenotypical correlations (Pearson’s R, bottom) and genetic correlations (rg, top) between latent factors and traditional ECG features.

Each LF was visualised by modifying that specific LF (at a range of ±1-3 SD of the LF mean as calculated in the BIDMC test set), while holding the others at their mean value (latent traversal, Fig. 2). The latent traversal indicated that different LFs were representative for independent ECG traits. This was further confirmed by the lack of correlation between the individual LFs (Fig. 2B), with only two LF showing an absolute Pearson correlation coefficient > 0.3 (LF 7 and 9, R = 0.31). Full 8-lead latent traversals, as well as an interactive plot of the LFs, are available in the Supplementary Materials.

**Figure 2:**
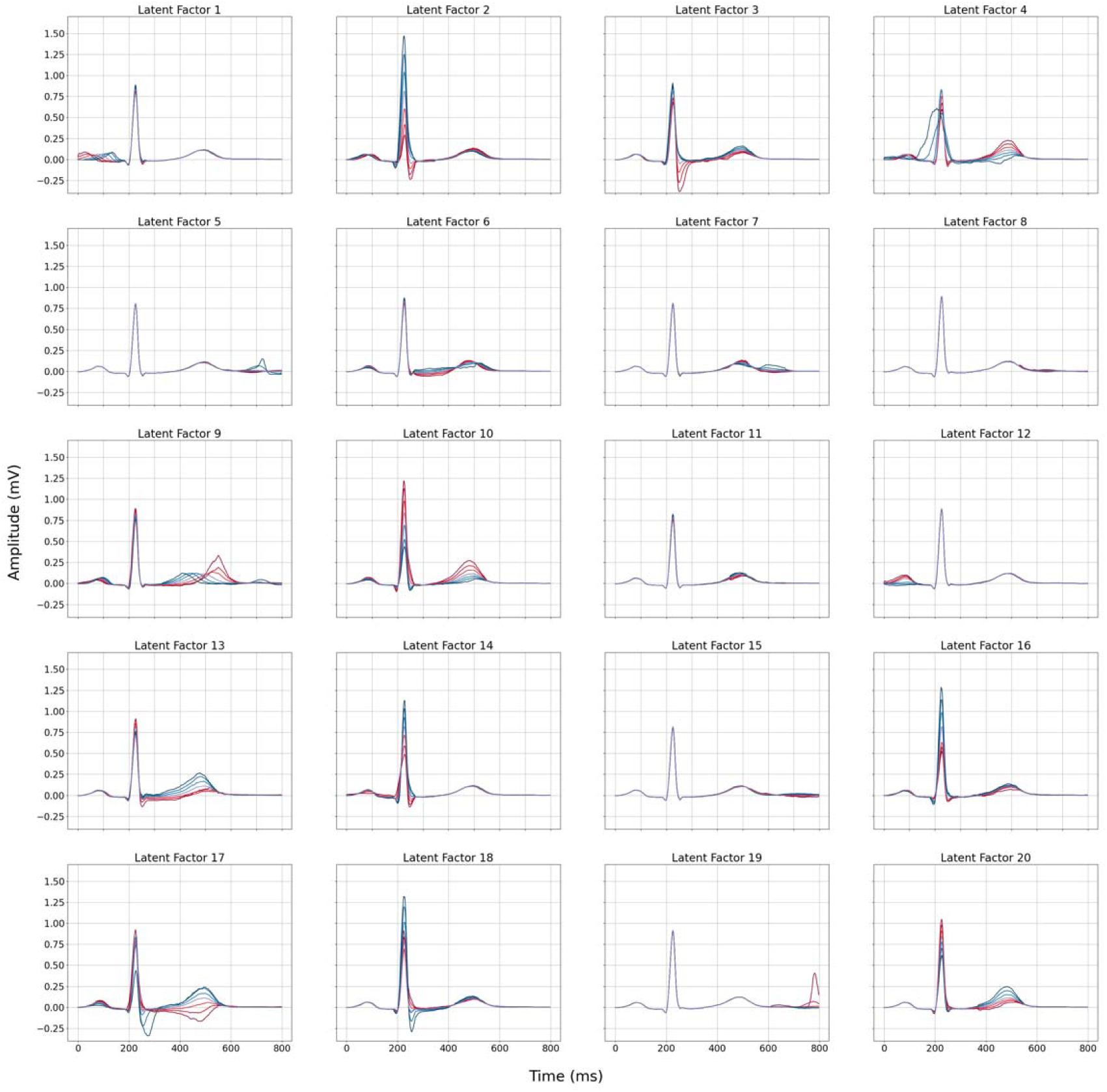
Lead I latent traversal plots of the 20 latent factors identified by the Variational Autoencoder demonstrate independent encoding of ECG components. Each plot corresponds to the reconstruction of the median beat in Lead I, exploring the influence of a single latent factor (range: -3 to +3 standard deviations from the mean), while maintaining others at the mean value. Red lines indicate negative deviations, while blue lines indicate positive deviations.

To explore the relationship between the novel LFs and traditional ECG parameters, we calculated the Pearson correlation coefficient between the LFs and the available ECG metrics in the UKB (P axis, PQ interval, QT interval, QTc interval, QRS duration, ventricular rate, PP interval, number of QRS complexes during the 10-second ECG (QRS number), P wave duration, R axis and T axis).

Most of the LFs capture information not represented in the traditional ECG features, as evidenced by their weak correlation with traditional ECG parameters, with only seven LFs displaying an absolute Pearson correlation coefficient above 0.3 with any ECG parameter (LFs 1-3 and 6-9, figure 1C). These seven LFs accounted for most of the associations with the traditional ECG parameters, though three of these ECG features (P-axis, P wave duration, T-axis) displayed a low (Pearson R < 0.3) correlation with the LFs. The highest correlation was observed for the QT interval with LF9 and the PQ interval with LF1 (Pearson R 0.78 and 0.77, respectively).

### The genetic architecture of the VAE latent factors

Overall, the LFs showed a variable degree of heritability, ranging from 3 to 20% (Supplementary Materials). Despite the lack of phenotypical correlation, several LFs proved to be genetically correlated with each other (absolute R between 0.0003 and 0.79; Figure 1B). Similarly, we observed higher values for genetic correlations with the traditional ECG features then the phenotypical correlations. Only three LFs showed a pairwise genetic correlation coefficient of less then 0.3. All genetic correlations between ECG parameters and LFs were above 0.3 (Figure 1C). These findings are indicative of the presence of pleiotropic genetic factors which act on multiple phenotypically uncorrelated ECG features. The associations of specific loci with ECG morphology are shown through locus specific LF traversals in the Supplementary Materials.

GWAS analysis of the 20 LFs identified 120 conditionally independent SNPs, corresponding to 118 genomic regions (P < 2.5 × 10^−9^) (Figure 3, Supplementary Materials). After combining regions with overlapping borders, we identified 65 unique loci that were associated with one or more LF. Prioritized genes were mapped to the GWAS regions by a combination of similarity based (polygenic priority score or PoPS)^10^ and locus-based (variant-to-gene or V2G)^11^ approaches, yielding 85 genes.^12^ The LFs were able to capture more oignificantly associated genes when compared to analysis of traditional ECG parameters on the same dataset. Traditional parameters were associated with 93 conditionally independent SNPs, corresponding to 51 unique regions that were associated (P < 4.5 × 10^−9^) with one or more traditional ECG parameter. Twenty-seven (42%) of the LF loci were not detected by GWAS of ECG parameters, while 13 (25%) of the regions identified by analysis of ECG parameters were not identified by the LF.

**Figure 3:**
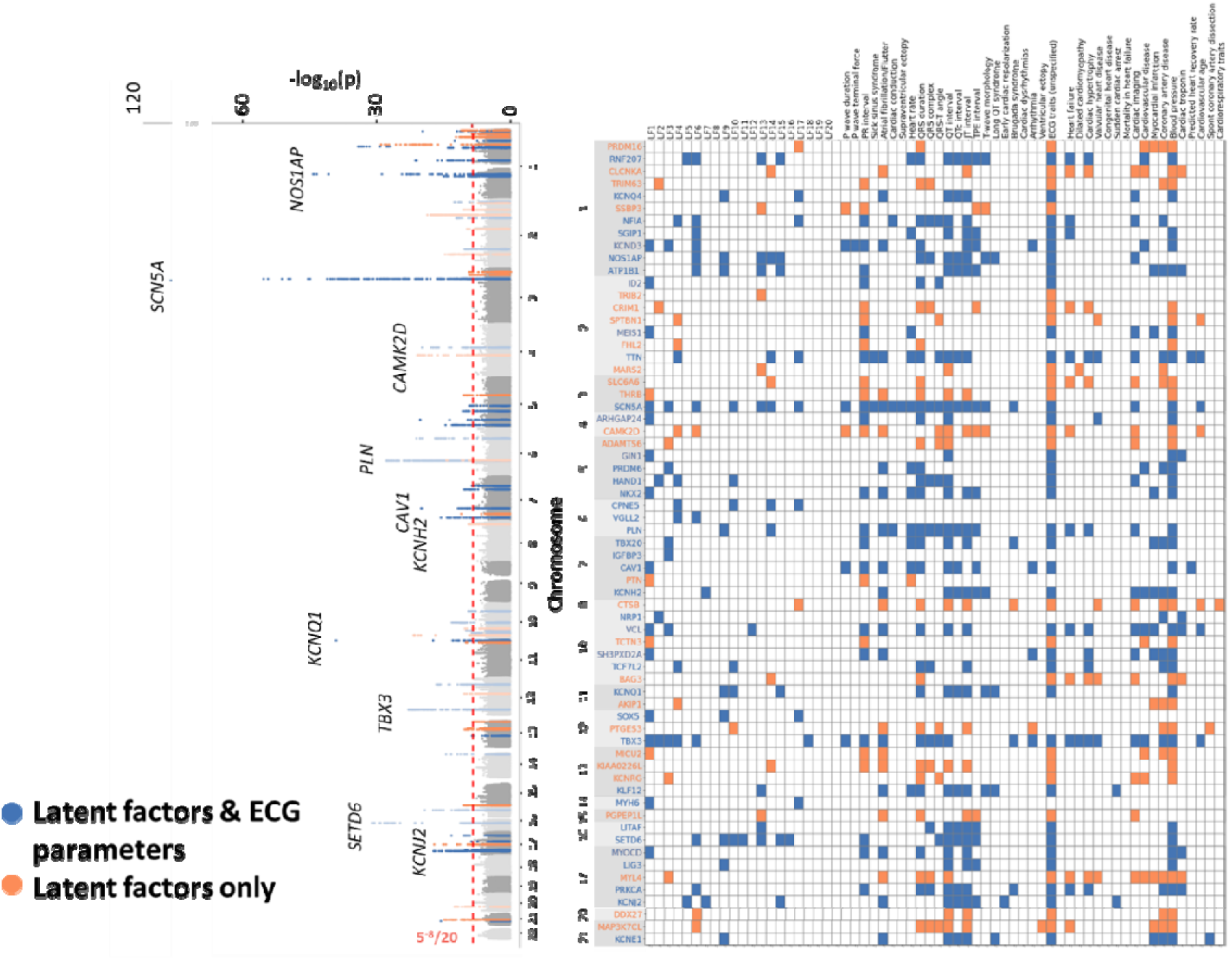
Latent factor GWAS joint Manhattan plot, with prioritized genes and their relationship to previous GWAS hits for ECG parameters and cardiac function related traits. Left side: the Manhattan plot collapses the latent factor (LF) traits into one plot by displaying the lowest p-value among the 20 LF GWAS for each of the included SNPs, calculated in the UK biobank. Right side: First listed prioritized gene (full gene list is available in the supplement) with their associated LF, previously reported ECG features, and cardiac function-related traits. The colours represent the comparison between the LF GWAS and traditional ECG parameters GWAS (including P axis, PQ interval, QTc interval, QT interval, QRS duration, Ventricular Rate, PP interval, QRS number, P wave duration, R axis and T axis). Blue: regions which are identified by the LF GWAS and the ECG parameter GWAS in our study and have been previously validated in other GWAS analyses of ECG traits. Orange: regions which are identified by LF GWAS only in our study (not conventional ECG traits), but have been previously validated in other GWAS analyses of ECG traits.

### Identification and validation of novel genomic regions associated with the ECG

In order to identify novel common variant associations with the ECG, we identified LF GWAS loci not linked to any ECG traits in the GWAS catalog. One locus (mapped to *NRP1*) qualified. For discovery purposes, we additionally evaluated a broader set of genes at a more relaxed threshold of <1% FDR. We identified 46 loci which associated with any of the LF and were not previously linked to the ECG. We compensated for the increased risk of false positive associations due to the relaxed significance threshold by reanalysing these candidate SNPs in a separate tranche of data from 18,987 UKB participants, which was released after the main analysis. We identified six SNPs which were directionally concordant and significant (p-value < 0.05) in the validation tranche. These loci_were mapped to the genes *NRP1, TRIOBP, EFEMP1, NEDD9, GPC6* and *SEC14L4.* Two of these six loci, mapped to *NRP1* and *NEDD9*, were also associated with traditional ECG parameters at the same significance threshold.

### Pathway analysis

We performed gene function analysis for all the prioritized GWAS genes (n = 85) with Functional Mapping and Annotation of Genome-Wide Association Studies (FUMA) software^13^. All protein-coding genes were considered as the background genes. The prioritized genes were more highly expressed in the heart (P 2.16 × 10^−8^) and blood vessels (P_adj_ 0.001). The analysis of gene-set enrichment, focusing on cellular components Gene Ontology terms, revealed 28 terms that exhibited significant enrichment within our gene list (Figure 4). Most of these terms were related to cardiac ion channels, the sarcomere, and the cytoskeleton.

**Fig 4:**
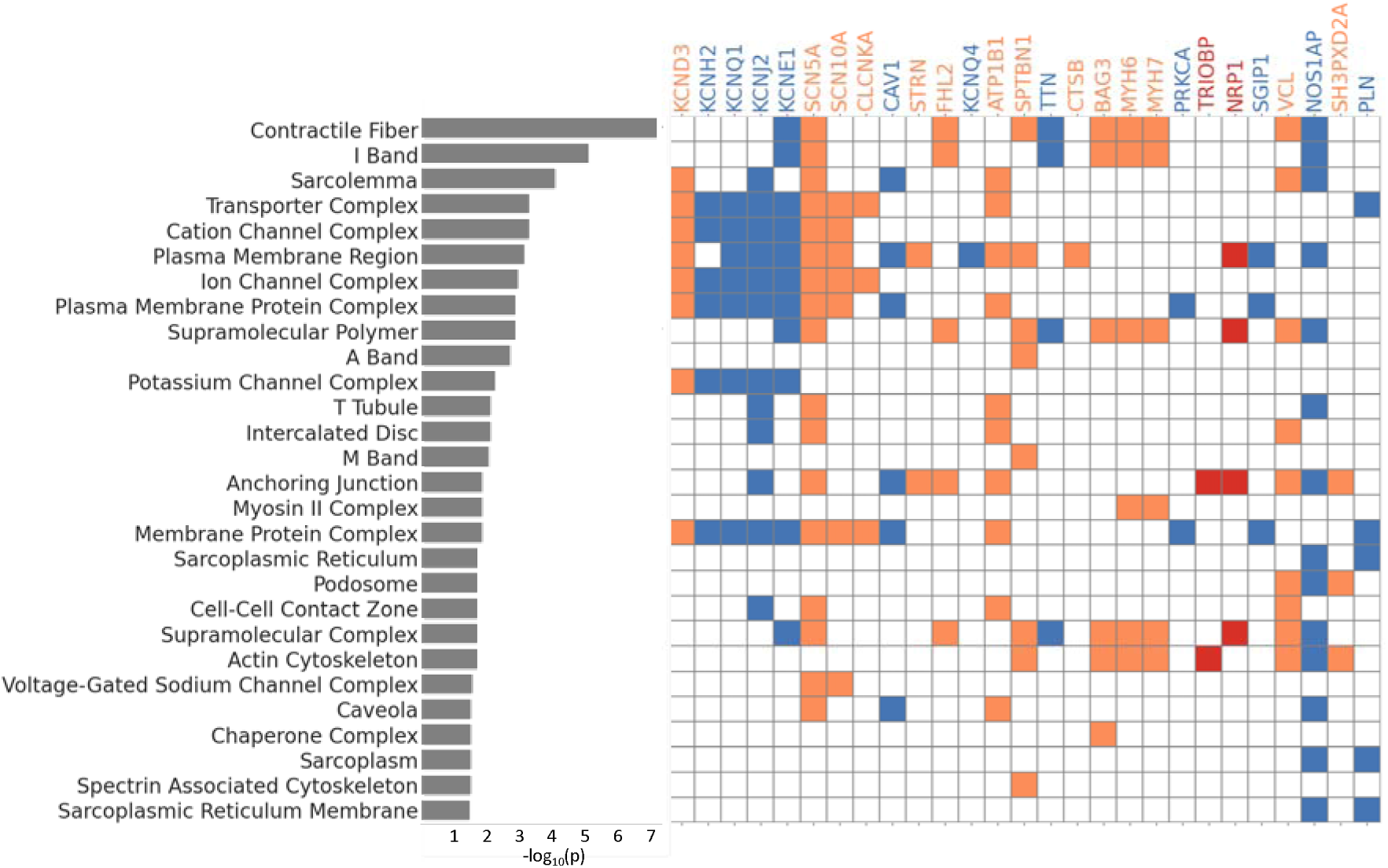
Significantly enriched Gene Ontology Cellular Components terms and the associated prioritized genes in the latent factor (LF) GWAS. Analysis of enriched pathways demonstrates shared molecular functions for genes identified by analysis of both traditional ECG parameters and LF, as well as the LF only and our novel findings. Blue: regions which are identified by the LF GWAS and the ECG parameter GWAS in our study and have been previously validated in other GWAS analyses of ECG traits. Orange: regions which are identified by LF GWAS only in our study and have been previously validated in other GWAS analyses of ECG traits. Red: novel regions, which have not been linked to the ECG in previous GWAS but have a p-value <1% in the LF GWAS and replicate in a held our validation tranche

### Gene-wise rare variant association study reveals additional genes associated with latent factors, and provides support for LF GWAS loci

The rare variant analysis was performed by gene burden testing, initially through an exome-wide approach, with subsequent subset analysis focusing on the prioritized genes from the GWAS (n = 85). With the whole-exome approach, we identified two genes, which demonstrated significant rare-variant associations with the LFs: *NEK6* with LF19 and *IL17RA* with LF5 (both for singleton protein-altering variants, Fig 5). Subsequent subset analysis focusing on the prioritized genes identified five additional associations (Fig. 5). Singleton variants in the *NME7* gene were associated with LF7. LF10 was associated with variants in *MYBPC3* with an allele frequency <0.001. Rare (allele frequency <0.001) variants in *CCT8* were associated with changes in LF11. Surprisingly, in the GWAS analysis, this locus was associated with LF7, rather than LF11. Low frequency (<0.01) variants in the *ADAMTS6* gene were associated with LF3. Finally, variants in *SCN5A* (<0.01 allele frequency) were associated with LF1 (Fig. 5), which is closely correlated to the PQ interval.

**Fig 5.**
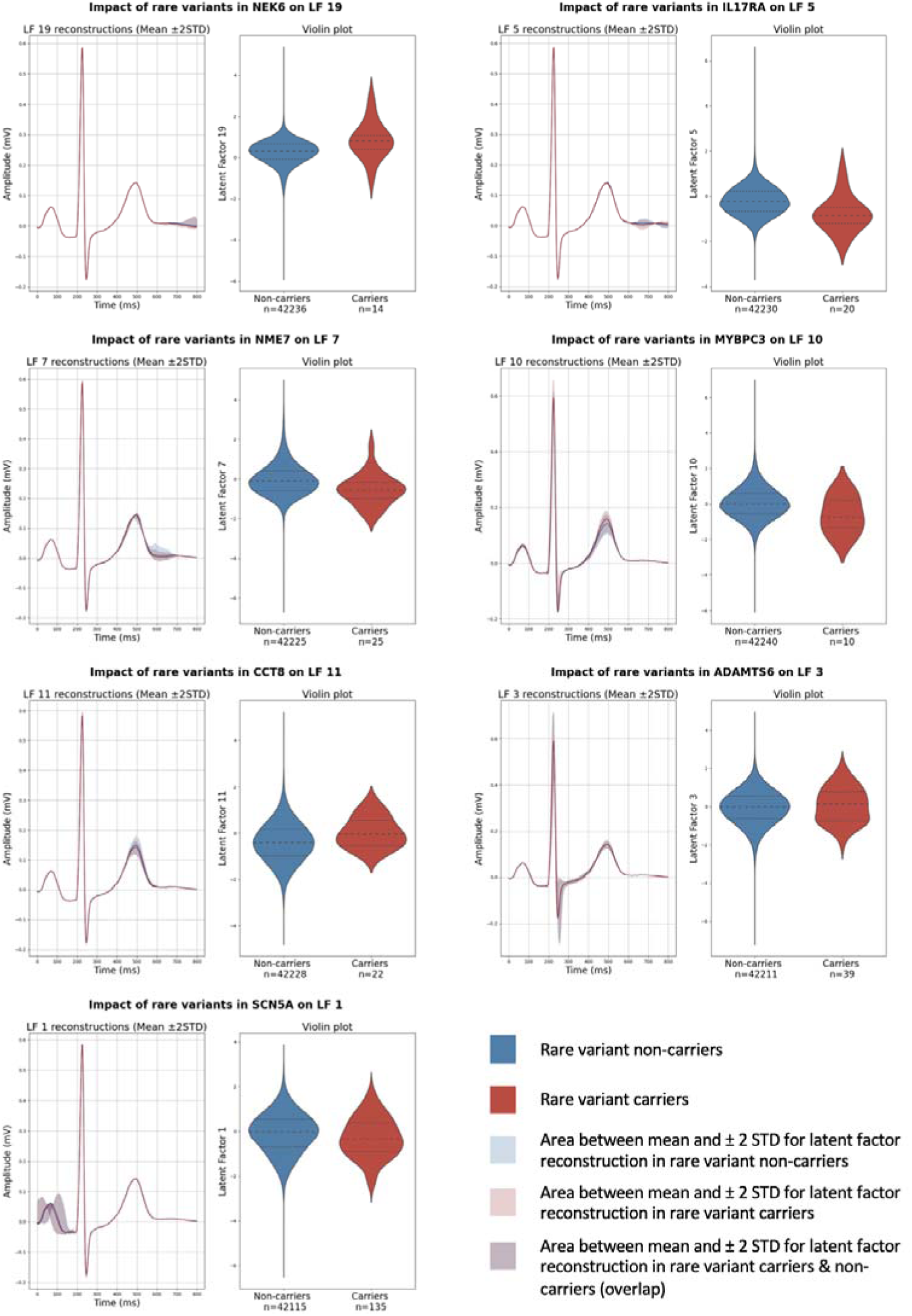
ECG median reconstructions and violin plots for carriers and non-carriers of rare variants associated with the latent factors. The median reconstructions were generated by holding all latent factors as a set value and altering the significantly associated latent factor. Dashed lines: medians, dotted lines: quartiles. The lines represent the reconstructions at the population mean and the shaded area indicating ±2 standard deviations.

### Phenotypic associations with the latent factors

To identify phenotypic associations, we developed separate regression models with the 20 LFs or the available ECG parameters as predictors and age, age^2^, and sex as covariates. The models were used to obtain phenotype predictions in the test set, which were correlated with the actual phenotype values (biserial correlation coefficient for binary traits and Pearson’s correlation coefficient for the continuous traits). We tested the correlation for 2074 binary disease phecodes in the BIDMC, 2093 continuous phenotype measures in the UKB and 39 echocardiographic traits in the BIDMC.

Overall, 798 disease phecodes were significantly correlated to at least one of the model predictions. The LFs identified 147 phenotypic correlations which were not significant for the ECG traits at the Bonferroni selection threshold (Fig. 6A), whereas 25 of the correlated traits identified by the ECG parameters were missed by the LFs. For phecodes correlated to both LF and ECG traits predictions, 76% (473/626) had a higher correlation coefficient with the LF. Although this trend was observed for all phecode categories, it was most pronounced for cardiac diseases (Fig. 6A). The ECG traits models were mainly driven by the ventricular rate (Fig. 6B), while the contributions of the LFs appeared more varied and category dependent.

**Fig. 6.**
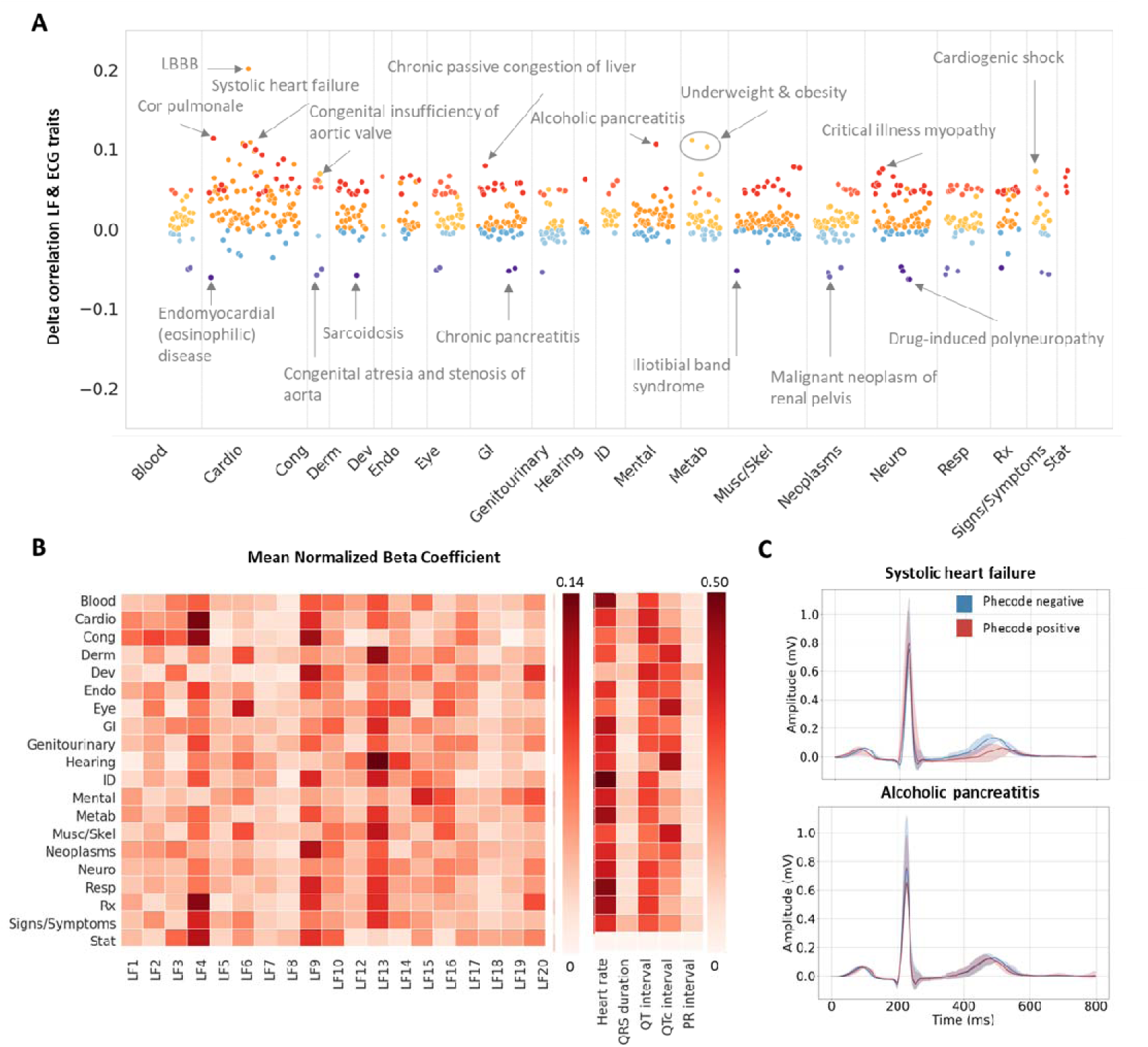
Latent factor and ECG trait associations with categorical disease labels (phecodes) in the BIDMC dataset. A: Difference between the point biserial correlation coefficients for the actual and predicted phecode status with the model trained on latent factors (LFs) or ECG traits. Positive deflections: higher correlations to the LF predictions, negative deflections: higher correlations to the ECG traits predictions. Colour code: red: phecodes only correlated to the LF predictions, orange: higher correlations with the LF model, blue: higher correlations with the ECG traits model, purple: phecodes only correlated to the ECG trait predictions. B: Heatmap of the category mean of the absolute beta parameter from the multivariate regression model, normalized to the sum of all beta coefficients for the category. C: The lead I median ECG reconstructions stratified by phecode status for systolic heart failureand alcoholic pancreatits. Lines: reconstructions at the population mean, shaded area: ±0.5 standard deviations. Category label abbreviations: Blood: Blood/Immune disorders, Cardio: Cardiovascular disorders, Cong: Congenital disorders, Derm: Dermatological disorders, Neoplasms, Neuro: Neurological disorders, Resp: Respiratory disorders, ID: Infectious diseases, Rx: Disorders due to external agents (e.g., medications), Musc/Skel: Musculoskeletal disorders, Mental: Mental health disorders, GI: Gastrointestinal Disorders, Metab: Metabolic Disorders, Derm: Dermatological Disorders, Endo: Endocrine Disorders, Dev: Developmental Disorders, Cong: Congenital Disorders, Stat: Status/Other

Similar findings were observed for the continuous phenotypes in the UKB (Fig 7A). Out of 1083 phenotypes which were significantly correlated to at least one of the model predictions, 158 were only identified with the LFs, whereas 20 were only correlated to the ECG traits predictions. Out of the remaining phenotypes, 58% (522/905) were more highly correlated with the LF predictions. Both heart rate and the QT(c) interval proved to be the main contributors to the ECG trait models across most categories (Fig. 7B).

**Fig. 7.**
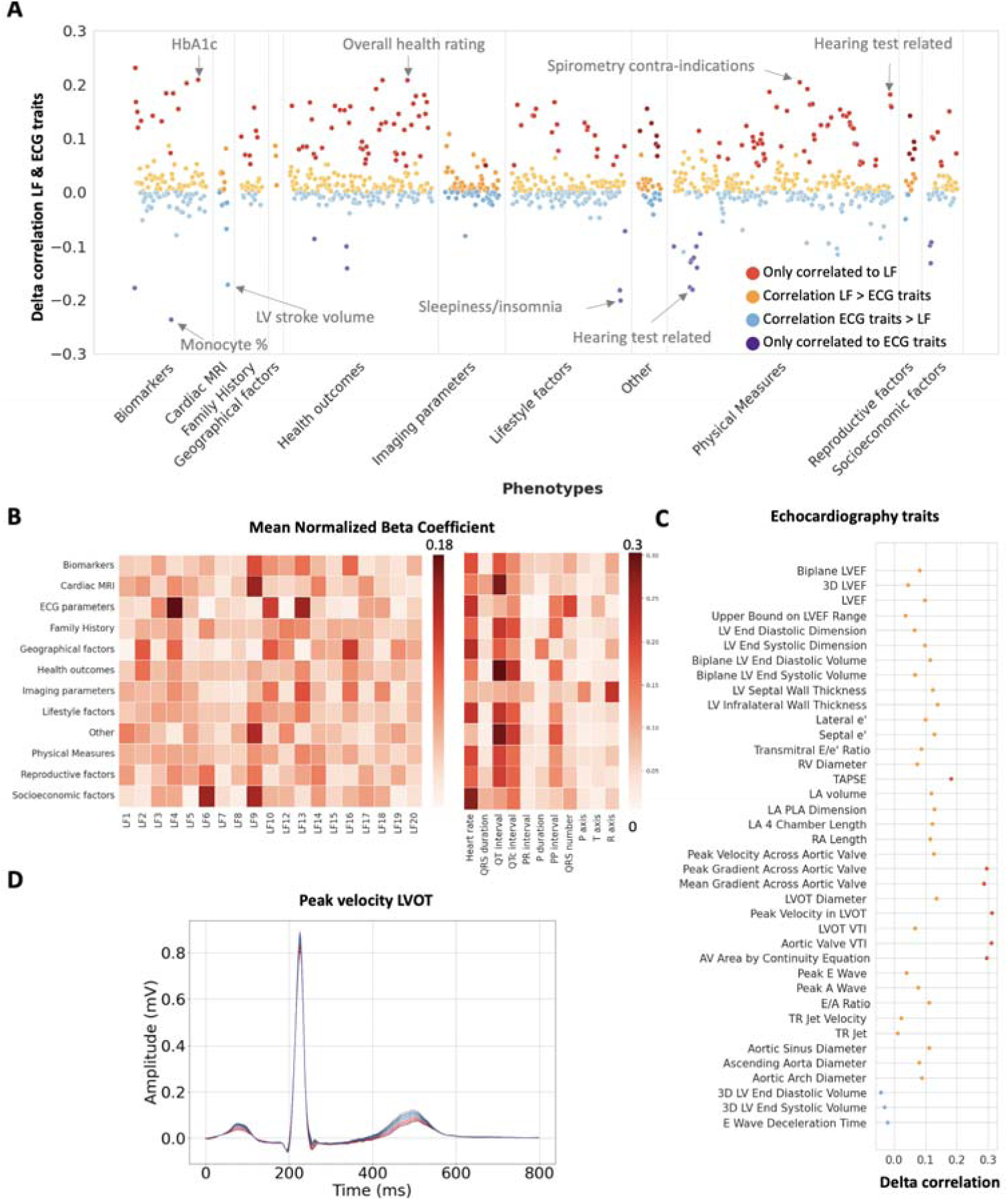
Latent factor and ECG trait associations with UK biobank quantitative phenotypes and echocardiographic traits in the BIDMC. A: Difference between the Pearson correlation coefficients for the actual and predicted continuous phenotypes in the UKB with the model trained on latent factors (LFs) or ECG traits. Positive deflections: higher correlations to the LF predictions, negative deflections: higher correlations to the ECG traits predictions. B: Heatmap of the category mean of the absolute beta parameter from the multivariate regression model, normalized to the sum of all beta coefficients for the category. C: Difference between the Pearson correlation coefficients for the actual and predicted echocardiographic traits in the BIDMC with the model trained on latent factors (LFs) or ECG traits. Colour code (A&C): red: phecodes only correlated to the LF predictions, orange: higher correlations with the LF model, blue: higher correlations with the ECG traits model, purple: phecodes only correlated to the ECG trait predictions. D: The lead I median ECG reconstructions based on the mean ±1-3 standard deviations values of left ventricle outflow tract (LVOT) peak velocity. Red lines indicate negative deviations, while blue lines indicate positive deviations.

The difference between the LF and ECG traits associations was most pronounced for the echocardiographic traits (Fig. 7C). We found that all 35 out of the 38 traits were more highly correlated to the LF predictions, with six traits only identified by the LF model and none only by the ECG traits. The advantage of the LF model was most pronounced for traits associated with the aortic valve and the left ventricular outflow tract (Fig. 7C-D).

## Discussion

Traditionally, the interpretation of the ECG relies on the identification and measurement of human defined morphologies and intervals. While the study of these conventional ECG parameters has yielded much insight into cardiac physiology and pathophysiology, these methods fail to fully capture the complexity of the ECG. The VAE model offers a novel approach which extracts features that more fully characterise the ECG, without anchoring on previous knowledge and concepts, and preserve high interpretability through direct signal visualisation.

Our VAE model was trained on a large (> 1 million ECGs) secondary care dataset, derived from both in- and outpatient settings, to capture a broad range of pathological and normal ECGs. By use of the open-source BRAVEHEART software, we provide a median beat extraction method which is easily transferable, as it does not rely on proprietary algorithms and can be applied to different types of digital ECG file formats. Additionally, our model was able to capture a high degree of electrophysiological variability while preserving feature disentanglement. This was demonstrated in external validation in the UKB, where our model achieved a Pearson correlation coefficient of 0.95, surpassing the 0.88 obtained by previous models^6^.

By examining the correlations and genetic associations of the 20 VAE-derived LFs and the 11 conventional ECG parameters, we were able to show that the LFs capture the majority of the of the phenotypical and genetic aspects of the conventional metrics. Additionally, the LFs were able to identify additional ECG characteristics and genetic associations. In a head-to-head comparison on a single dataset, while most genetic associations with the conventional ECG parameters were captured by the LF, 52% of the LF hits eluded detection by the ECG parameters.

Overall, we observed lower correlation coefficients between the LF themselves and the LFs and conventional ECG features, compared to their genetic correlation. Many genetic loci were found to associate with multiple phenotypically uncorrelated ECG features. These findings are indicative of the presence of pleiotropic genetic factors which act on multiple apparently unrelated ECG features.

The LF were able to capture genetic associations with many well known modifiers of cardiac electrophysiology (e.g. *SCN5A*, *KCNQ1*, *NOS1AP*, *KCNH2*) as well as myocardial structure and contractile function (e.g. *TTN*, *MYH6*). Moreover, our study expanded the genetic landscape of ECG associations by identifying and validating six GWAS loci not previously linked to the ECG. Two of the the six prioritized genes in these loci (*NRP1* and *TRIOBP*) are associated with the same cellular component ontology terms as the previously validated GWAS genes from our study (e.g., *KCNQ1, SCN5A* and *MYH6-7*). Five of the six novel genes have previously been implicated in diverse cardiovascular processes. These novel findings may guide further mechanistic and therapeutic exploration into these genes. The *NRP1* gene (Neuropilin-1) has been linked to cardiac regeneration in zebrafish^14^. The *TRIOBP* (TRIO and F-actin Binding Protein) gene is involved in the modulation of the assembly of the actin cytoskeleton^15^. *TRIOBP*-*1*, a splicing isoform of *TRIOBP* has been previously shown to interact with the cardiac potassium channel *KCNH2* ^16^, with a potential effect on cardiac repolarization. In our study, *TRIOBP* associated with LF1, which is closely associated to the PQ-interval, potentially identifying a novel role in cardiac electrophysiology for this gene.

The *EFEMP1* gene encodes fibulin-3, an extracellular membrane glycoprotein, expressed in many human tissues including the heart^17^. This gene is upregulated in heart failure and recent studies in murine models have demonstrated a role in cardiac remodelling following ischaemic injury^18^. Neural precursor cell expressed developmentally down-regulated 9 (*NEDD9*) has been previously linked to endothelial fibrosis and pulmonary arterial hypertension^19^. *GPC6*, a member of the glypicans family of evolutionary conserved heparan sulfate proteoglycans anchored to the extracellular leaflet of the cell membrane, has been associated with heart failure in clinical and murine studies^20^.

Apart from the common variant associations, we were also able to identify seven genes with rare variant based associations with the LFs. Two of these (*MYBPC3* and *SCN5A*) are well-established contributors to cardiomyopathy^21^ and cardiac electrophysiology^22^. The ‘never in mitosis A’ related kinase 6 (*NEK6*) gene has been previously linked to hypertrophic cardiomyopathy (HCM)^23^ and cardiac fibrosis^24^ in mouse models, but had not previously been connected to human disease or the ECG. The interleukin 17 receptor A gene (*IL17RA*) encodes a membrane glycoprotein that binds proinflammatory cytokines. *IL17RA* has also been associated to inflammatory dilated cardiomyopathy ^25^ and heart failure^26^.

The *NME7* gene belongs to a family of nucleoside diphosphate kinases with a role in ciliary transport. Common variants in the *NME7* locus have been previously linked to the QT interval^27^, although the causal gene had not yet been identified. The *CCT8* gene encodes a subunit of the chaperone complex involved in folding of actin and tubulin^28^. Common variants in the *CCT8* locus have recently been linked to HCM^29^. The *ADAMTS6* gene encodes a protease, involved in the regulation of extracellular matrix composition^30^. Coding variants in the *ADAMTS6* gene have been linked to the QRS interval^31^.

Apart from genomic associations, the VAE-derived features also proved a more powerful tool for the identification of phenotypic associations, especially for cardiac disease phecodes and echocardiographic traits. For these analyses the LF both enabled the discovery of novel associations, otherwise overlooked by conventional ECG features, and boosted the power of known association testing. Additionally, we demonstrated that LF visualisations can provide a clear indication of the ECG changes associated with the relevant genotypes and phenotypes. The ability to easily visualise the genetic and phenotypic associations (also made available interactively at https://www.cardiodb.org/decg_explorer/) highlights the potential of the VAE approach for the generation of new hypotheses. This approach can facilitate the discovery of novel phenotypes and guide further genetic and mechanistic explorations.

In conclusion, our study addresses limitations of traditional ECG interpretation by introducing a novel approach using the VAE. Our model, trained on a large, diverse dataset, enhances interpretability, but also surpasses previous methods in capturing variability and identifying associations for common and rare-variant genetic variants and phenotypical traits. These findings contribute valuable insights into cardiac electrophysiology, emphasizing the potential of advanced analytical methods like the VAE for unravelling the complexity of the ECG.

## Methods

### Dataset

The VAE was trained and validated on a de-identified dataset derived from the Beth Israel Deaconess Medical Center (BIDMC dataset). This dataset consists of 1,169,387 resting 10 second ECGs derived from 189,542 patients examined at the hospital (median age 65 years ± 16 SD at the time of examination). The BIDMC dataset is supplemented by dated ICD codes and tabular echocardiographic data.

The VAE model was externally validated in the UK Biobank (UKB). The UKB is a prospective study of 40-69-year old volunteers from the general population, recruited between 2006 and 2010. Digital ECG recordings were taken during the imaging visits (instance 2). Extensive phenotypic and genetic data has been gathered from the participants and were accessed with approvals in place (Application numbers 47602 and 48666)^32^. Genotyping was performed by the UKB central team with the Applied Biosystems (Affymetrix) UK BiLEVE Axiom Array or the UKB AxiomTM Array^32^. Imputation was done with the Haplotype Reference Consortium and the merged UK10K and 1000 Genomes phase 3 (1000G) reference panels^32^. Data used in this study were from a subset of UKB participants for whom ECGs were available (n = 42,248).

### Median ECG derivation

All median beat ECGs were obtained from resting 10-second ECG signals with BRAVEHEART ECG analysis software (https://github.com/BIVectors/BRAVEHEART)^9^. The median beat was further adjusted by cropping or zero-padding to a length of 0.8 sec (sampled at 500 Hz) and aligning the signals by cross-correlation. In order to remove signals with large pacing spikes or excessive noise, signals with extreme voltages (<-8 mV or >8 mV) were excluded from the analysis. After median ECG derivation and filtering, the remaining 1,165,311 median ECGs were randomly split into a training set (1,048,778 ECGs from 180,679 individuals), a validation set (58,265 ECGs from 4,233 individuals) and a test set (58,268 ECGs from 4,500 individuals), in a 90/5/5% split.

### VAE architecture

The VAE consists of three components: the encoder, the decoder and the latent space. The encoder and the decoder are made up of one-dimensional convolutional layers with increasing filters and decreasing kernel sizes closer to the latent space. The latent space was restricted to 30 features at a maximum, although typically only a subset of these features was used by the model for the reconstruction. The model was trained to minimize both the median ECG reconstruction loss, defined by a symmetric mean absolute percentage error function, and the Kullback-Leibler divergence (KL loss). This second term is specifically added to the VAE model to ensure that the features generated by the model are generative and disentangled. An additional β-parameter was included as a weight on the KL-term to optimize the balance between the reconstruction loss and the latent factor interpretability. We tested beta values of 0.1, 0.25, 0.5, 1, 3, 5 and 10 and defined the best model at a β-parameter of 0.25 based on the Pearson correlation between the median and its reconstruction in the validation dataset, as well as a visual inspection of the latent vector traversals.

### GWAS and common variant gene analysis

The UKB participants included in the genetic analyses were selected for European ancestry, missingness rate of SNPs <10%, no sex discrepancies, and removing outliers of heterozygosity or relatedness. After selection, ECGs were available for 31,118 individuals. Quality control was performed to exclude SNPs with a minor allele frequency <0.1%, genotyping rate <95%, deviation of heterozygosity with Hardy-Weinberg equilibrium p < 1.0 × 10^−8^ or <0.4 INFO imputation score.

The GWAS was carried out with the FastGWA MLM implemented by the Genome-wide Complex Trait Analysis (GCTA) software using a genetic relationship matrix (GRM) to adjust for population structure^33^. The latent factor distributions were normalized by rank-based inverse normal transform prior to the analysis. Age, sex, height, BMI, the UKB assessment centre and the first 10 genetic principal components were included as covariates. We report both the SNPs which were identified by the conventional genome-wide significance threshold, with Bonferroni correction for testing of multiple features (p-value < 5 × 10^−8^/20 for the LF and < 5 × 10^−8^/11 for the traditional ECG parameters). A less stringent threshold at <1% false discovery rate (FDR) was applied to the LF GWAS to select loci which were not previously associated with the ECG in the GWAS catalog.

The genetic variance explained by genome-wide SNPs (SNP-based heritability) was calculated with the genomic-relatedness-based restricted maximum likelihood (GREML) analysis using the GCTA software^34^. Genetic correlation was calculated with the bivariate GREML analysis method^35^.

The common variant gene analysis was performed with MAGMA software on the summary statistics obtained from the GWAS^12^. We used 18,383 genes with a genome-wide significance threshold defined as P=2.72 × 10^−6^.

### Locus identification and finemapping

Conditionally independent genetic variants were identified using a chromosome-wide stepwise conditional-joint analysis implemented in the GCTA software^36^. Variants within 500 kb of each other were aggregated and an additional 500 kb were added to flank the variants on each side of the locus. A locus was considered novel if it was not previously associated to electrocardiographic traits according to the GWAS catalogue. Potential novel loci were further validated by a literature search of nearby genes.

Functionally informed fine mapping with PolyFun^37^ and SuSiE^38^ was performed to identify the most likely causal variants. Precomputed prior causal probabilities from the PolyFun UKB analysis were used to compute the per-SNP heritability. Linkage disequilibrium was calculated for each locus and used for finemapping. Using SuSiE, we calculated the per-SNP posterior inclusion probability (PIP) and identified 95%-credible sets of likely causal variants, under the assumption of at most 5 causal variants for each locus. These variants were used for annotation with the nearest protein-coding gene and gene prioritization. As several regions did not contain 95%-credible sets of likely causal variants for the regions identified by the 5% FDR threshold, SNPs with the lowest p-values were used instead.

### Gene prioritization

Candidate genes were selected using three approaches: nearest gene annotation, the polygenic priority score (PoPS)^10^ and variant-to-gene (V2G)^11^. PoPS uses gene expression, biological pathways and protein-protein interactions to assign priority to genes, based on their similarity to other potentially causal genes identified based on a prior functional annotation and gene-based analysis of the GWAS summary statistics by the MAGMA software.^12^ Genes outside the GWAS loci which were significantly associated with the phenotype according to the MAGMA analysis were reported separately. The V2G method is based on the integration of epigenomic data (eQTL, pQTL, sQTS, chromatin interactions), functional predictions and genomic distance to assign a variant level score, with higher scores representing a higher likelihood of a functional effect on a target gene. The candidate list was generated by selecting the top three highest scoring genes for both the PoPS and V2G methods, as well as the MAGMA significant genes and the nearest protein coding gene for each region (Supplementary Materials).

To prioritise the most likely causal gene for each locus, we first selected all established Mendelian cardiac arrhythmia^39^ and cardiomyopathy^40^ genes out of the candidate list. Secondly, we selected the genes which were selected by >1 method (PoPS, V2G and MAGMA). If several genes were selected by an equal number of methods at one locus, a literature search was conducted to identify the gene with the strongest relationship to cardiac function. If multiple genes were plausible, we selected all candidates for the region. If there was no evidence for any of the genes in literature, we selected the nearest gene.

### Pathway analysis

The GWAS prioritized genes were assessed for tissue specificity in the GTEx v7 (30 general tissue types) and gene set enrichment for molecular function Gene Ontology terms with all protein coding genes as background with Functional Mapping and Annotation of Genome-Wide Association Studies (FUMA) software^13^.

### Validation of novel GWAS hits

We performed GWAS validation in a separate cohort of UKB digital ECGs collected more recently, which became available at time of writing this manuscript. This dataset consists of ECGs from 24,355 participants, 18,987 of which were included in the GWAS validation cohort after quality control. As the lower sample size resulted in a lack of model convergence for some LF with the fastGWA-REML (GCTA) analysis, the validation cohort GWAS was done with REGENIE software^41^. Variants from the LF GWAS significant at the FDR <1% threshold, which were not previously associated with ECG traits, were selected for validation. Variants with a p-value of less then 0.05 in the validation cohort were reported as novel findings.

### Rare-variant gene-based association testing

We performed rare-variant gene-based association testing for each normalized latent factor with the UKB 500k whole-exome sequencing (WES) data using the REGENIE software^41^. As a first step, a regression model was fitted to correct for polygenicity, ancestry and relatedness, with the SNPs which were previously selected for the GWAS analysis. In the second step, we preformed exome wide gene-based burden testing, with a significance threshold defined at false discovery rate (FDR) < 5%. Covariate adjustment for sex, age, age^2^, height, BMI and the 10 first genetic principal components was included. Masks were constructed based on variant frequency (singleton, <0.001 and <0.01) and likelihood of a pathogenic effect. For missense variants, this was determined by a pathogenicity score based on a combination of five prediction tools (SIFT, PolyPhen2 HDIV, PolyPhen2 HVAR, LRT and MutationTaster)^42^. As previously published, variants which were predicted deleterious by all five algorithms were considered as “likely deleterious”^42^. Only loss-of-function and likely deleterious missense variants were included in the analysis. We initially considered 18,985 protein coding genes with subsequent subset analysis focusing on the prioritized genes from the GWAS analysis, with the FDR-corrected p-values adjusted to the number of genes for each analysis.

### Phenotypic associations

Phenotypic association studies were performed in the BIDMC for binary disease phecodes (n = 2074) and echocardiographic traits (n=39), and in the UKB for continuous phenotype traits (n = 2093). We developed separate regression models with the LFs or the available ECG parameters as predictors and age, age^2^, and sex as covariates. The BIDMC ECG parameters were heart rate, QRS duration, QT/QTc interval and PR interval, for the UKB P axis, PQ interval, QTc interval, QT interval, QRS duration, Ventricular Rate, PP interval, QRS number, P wave duration, R axis and T axis were used.

The models were used to obtain phenotype predictions, which were correlated with the actual phenotype values (biserial correlation coefficient for binary traits and Pearson’s correlation coefficient for continuous traits). Correlations with a Bonferroni corrected p-value below the 0.05 threshold were excluded from the analysis.

For the BIDMC dataset, the models were developed with ECGs from the VAE training dataset, predictions were derived from ECGs in the test set. We excluded phenotypes with >90% missing values and binary phenotypes with counts < 100 in the training set. For the disease phecodes we used a single random ECG per subject (n = 176,536 for training, 9,282 for predictions). For the echocardiographic traits we used a single ECG per subject, taken <30 days before or after the echocardiography (n = 58,063 for training, 3006 for predictions). For the UKB dataset, we randomly split the dataset into a training (n = 32,832) and a test set (n = 8208).

## Data availability and ethics

The summary statistics supporting the GWAS findings will be made publicly available through the GWAS Catalog upon publication following peer review. The code used to perform the analyses and generate the plots for this is accessible in the supplement. All UKB data used in this study is publicly availableto registered researchers (https://www.ukbiobank.ac.uk/). The LF generated from the UKB ECGs will be made available as a Returned Dataset in the UKB.

All studies were approved by the relevant regional research ethics committees, and adhered to the principles set out in the Declaration of Helsinki. The UK Biobank study was reviewed by the National Research Ethics Service (11/NW/0382, 21/NW/0157). This study was conducted under terms of access approval number 47602 and 48666. The BIDMC cohort ethics review and approval was provided by the Beth Israel Deaconess Medical Center Committee on Clinical Investigations, IRB protocol # 2023P000042. Access to the BIDMC dataset is restricted due to ethical limitations.

## Acknowledgments

The authors would also like to thank the InSIGHT Core in the Center for Healthcare Delivery Science at Beth Israel Deaconess Medical Center for assistance in obtaining primary data.

## Notes

**Sources of financial support:** This work was supported by Sir Jules Thorn Charitable Trust [21JTA], Medical Research Council (UK), British Heart Foundation [RE/18/4/34215; FS/CRTF/21/24183; RG/F/22/110078, FS/IPBSRF/22/27059], NIHR Imperial College Biomedical Research Centre, and an EJP RD Research Mobility Fellowship (European Reference Networks) to ES. AS is funded by a British Heart Foundation (BHF) clinical research training fellowship (FS/CRTF/21/24183). FSN and NSP are supported by the BHF (RG/F/22/110078 and RE/18/4/34215) and the National Institute for Health Research Imperial Biomedical Research Centre. LP is funded by a Medical Research Council (MRC) clinical research training fellowship (MR/Y000803/1). For the purpose of open access, the authors have applied a creative commons attribution (CC BY) licence to any author accepted manuscript version arising.

### Competing Interest Statement

JSW has received research support from Bristol Myers Squibb, and has acted as a consultant for MyoKardia, Pfizer, Foresite Labs, Health Lumen, and Tenaya Therapeutics. JWW has received research support from Anumama.

### Funding Statement

This work was supported by Sir Jules Thorn Charitable Trust [21JTA], Medical Research Council (UK), British Heart Foundation [RE/18/4/34215; FS/CRTF/21/24183; RG/F/22/110078, FS/IPBSRF/22/27059], NIHR Imperial College Biomedical Research Centre, and an EJP RD Research Mobility Fellowship (European Reference Networks) to ES. AS is funded by a British Heart Foundation (BHF) clinical research training fellowship (FS/CRTF/21/24183). FSN and NSP are supported by the BHF (RG/F/22/110078 and RE/18/4/34215) and the National Institute for Health Research Imperial Biomedical Research Centre. LP is funded by a Medical Research Council (MRC) clinical research training fellowship (MR/Y000803/1). For the purpose of open access, the authors have applied a creative commons attribution (CC BY) licence to any author accepted manuscript version arising.

